# Field evaluation of specificity and sensitivity of a standard SARS-CoV-2 antigen rapid diagnostic test: A prospective study at a teaching hospital in Northern Ghana

**DOI:** 10.1101/2021.06.03.21258300

**Authors:** Alhassan Abdul-Mumin, Abdulai Abubakari, Faith Agbozo, Abass Abdul-Karim, Benjamin Demah Nuertey, Kareem Mumuni, Anna-Katharina Heuschen, Lisa Hennig, Claudia M. Denkinger, Olaf Müller, Albrecht Jahn

**Author notes:** These authors contributed equally to the paper (Claudia M. Denkinger, Olaf Müller, Albrecht Jahn).

## Abstract

**Background:** The testing capacity for SARS-CoV-2 in Africa is rather limited. Antigen-detection rapid diagnostic tests (Ag-RDTs) are a cheap and rapid alternative to reverse transcriptase-polymerase chain reaction (RT-PCR) tests, but there is little data about their performance under real life conditions in tropical countries.

**Objective:** To evaluate the performance of a standard Ag-RDT in a population of a major hospital in northern Ghana.

**Methods:** Prospective, cross-sectional, blinded verification of the performance of the SD Biosensor Standard Q SARS-CoV-2 Ag-RDT under real life conditions in 135 symptomatic patients and 58 contacts of RT-PCR positives at Tamale Teaching Hospital in February 2021. Nasopharyngeal samples were taken under standard conditions and tested against RT-PCR in the hospital laboratory.

**Results:** 193 participants (median age 35 years, 109 male) were included into the study for which both RT-PCR test and Ag-RDT results were available. A total of 42 (22%) were RT-PCR positive. Of the 42 RT-PCR positives, 27 were Ag-RDT positive, resulting in a sensitivity of 64% (95% CI 49-79). Sensitivity among symptomatic patients was 58% (95% CI 38-78). 123 were identified Ag-RDT negatives of the 151 RT-PCR negatives, resulting in a specificity of 81% (95% CI 75-87).

**Conclusions:** SARS-CoV-2 Ag-RDTs appear to have a rather low sensitivity and particularly a low specificity under real life conditions in Africa. The role of existing Ag-RDTs in countries with high-temperature climates and limited resources still needs more data and discussion.

## Background

The emergence of COVID-19 in China by the end of 2019 has led to the largest pandemic in recent human history (1, 2). It had initially been predicted, that Africa would become the worst affected global region, due to its weak health systems, prevailing poverty, and the existing high burden of infectious diseases (3, 4). However, by the end of 2020, only some 2% of the global number of cases and deaths were reported from the WHO African Region, but there have recently been signs for a resurgence of the number of cases (5).

There are various potential reasons for the low number of cases reported from sub-Saharan Africa (SSA). The likely main reasons are a much lower testing capacity in most countries of SSA and a much younger population associated with fewer symptomatic cases; however, other factors such as climate, which may affect transmission dynamics, the effects of early public health response measures, herd immunity due to cross reactions with other corona viruses or prevailing parasitic infections may also play a role (4). Findings from SARS-CoV-2 seroprevalence surveys support the growing evidence of under-reporting and a high proportion of asymptomatic and mild cases in SSA countries (6). In Zambia for example, a population-based survey has shown that the number of laboratory-confirmed SARS-CoV-2 cases are underestimating the real degree of community transmission at least 100-fold (7). However, SARS-CoV-2 serology results in SSA resulting from commercial tests validated outside Africa need to be interpreted with caution (8).

Testing is essential for the diagnosis of COVID-19 patients and to identify those persons who are infectious. Molecular assays to diagnose SARS-CoV-2 are considered the gold standard for detection of SARS-CoV-2 infection. They are typically based on RT-PCR to detect viral RNA and are highly sensitive and specific (9). However, they require a good laboratory infrastructure, trained staff, expensive equipment, and results are usually available only with significant delay (10). Single use, lateral flow, antigen-detection rapid diagnostic tests (Ag-RDTs) are a cheaper and easy to use alternative to RT-PCR tests. They are considered as a useful supplement to RT-PCR testing, in particular as they mainly identify cases in the early phase of disease and with high viral load and thus likely to be infective, and as they provide results within 15 minutes (11, 12). An increasing number of Ag-RDTs has become authorized by the US Food and Drug Administration (FDA) and other health authorities, which have been shown to be highly specific, but not as sensitive as molecular tests (10). WHO recommends a minimum sensitivity of 80% and a minimum specificity of 97% for SARS-CoV-2 Ag-RDTs (10).

We here report the results of a field evaluation of a standard SARS-CoV-2 Ag-RDT (SD Biosensor Standard Q) in Ghana which assesses whether the performance reported in clinical studies in mostly high-resourced countries with moderate climates holds up in low-resourced settings with high-temperature climates.

## Methods

### Study location and population

This study was conducted in the Tamale Teaching Hospital, situated in Tamale in the Northern Region of Ghana. It serves as the only tertiary health care facility for the Northern Region (where Tamale is located), Northeast, Savannah, Upper East and Upper West Regions as well as parts of the Bono East and Oti Regions of Ghana. Together, the population of the catchment area is about six million. The hospital also serves as the clinical training setting for students of the *University for Development Studies*. Since the detection of the first case of COVID-19 in Ghana in March 2020, the hospital has been at the forefront of the fight against the pandemic in the northern sector of the country. It hosts the only treatment center for COVID-19 as well as the zonal laboratory, where most samples taken from suspected COVID-19 cases are tested.

### COVID-19 in Ghana

The first cases of COVID-19 in Ghana were recorded on March 12, 2020, while the first case in Tamale has occurred about two weeks after this. By September 9, 2021, Ghana – with a population of about 30 million - had a total of 123,874 reported SARS-CoV-2/COVID-19 cases and 1098 reported deaths (13). The Northern Region has recorded 1,713 cases to date, with the majority of these cases coming from the Tamale metropolis. A lockdown, mainly for Greater Accra and some parts of the Ashanti Region, was imposed for two weeks in April 2020 to stem the control of the virus spread. While the first epidemic wave lasted from March until November 2020, a second wave is currently under way, driven mainly by new strains of the virus with higher infectivity, and increasing rates of hospitalization due to severe symptoms (13, 14). Ghana has a limited capacity to run RT-PCR tests, which has resulted in significant delays in getting RT-PCR test results during the first and second waves of the pandemic.

### Study design and participants

This study is a cross-sectional, blinded verification study of the performance of the Standard Q SARS-CoV-2 Ag-RDT under real life conditions at the Tamale Teaching Hospital. From February 15 to 20, 135 consecutive patients and 58 contacts were recruited. The patients were eligible if they were referred by an attending physician because they exhibited signs suggestive of COVID-19 (symptomatic), or if they were contacts of patients who tested positive for COVID-19 by RT-PCR test. None of the asymptomatic participants reported back to hospital with symptoms within 14 days after the sample collection.

The sample size of 200 was determined using the Cochrane (1977) formula for sample size calculation.

Following verbal informed consent, clinical and demographic data were recorded from the study participants on a standard questionnaire, including specific symptoms, age, and sex. None of the participants declined for samples to be taken and to participate in the study.

### Index test

The Ag-RDT evaluated was the STANDARD Q SARS-CoV-2 Ag Test (SD Biosensor, Inc. Gyeonggi-do, Korea), which is distributed by Roche (15). The test was purchased in Switzerland and transported to the Institute of Global Health in Heidelberg, Germany, using standard procedures. From Heidelberg, the tests were sent to Tamale, Ghana, by air, again using standard procedures for transport of laboratory materials. In Tamale, the test kits were stored in a designated storage room at the Tamale Teaching Hospital at temperatures between 22°C and 27°C. Testing was done according to manufacturer’s instructions for use. However, both the COVID-19 isolation center and the general wards were not air-conditioned, and the environmental temperature fluctuated between 24°C and 37°C. Most of the tests were done during daytime when the temperature was highest.

### Reference test

Reverse transcriptase-polymerase chain reaction (RT-PCR) was used as the reference test. The RT-PCR samples were collected by health-care workers using nasopharyngeal swabs. At the study laboratory, the samples were initially lysed to inactivate any viral agent, followed by extraction of the viral RNA in separate biosafety class 2 cabinets. The next step involved the mastermix preparation and template addition in separate hoods. The final stage was amplification using AriaMx real-time PCR thermocycler. The reagent used was LightMix® SarbecoV E-gene (manufacturer TIB MOLBIOL) which is the standard assay used in the *Public Health Reference Laboratory*. The assay targets only the E-gene of SARS-CoV-2. Further details and test properties can be found in the instructions for use (16). A Ct value of <40 is interpreted as a positive and Ct values ≥40 as a negative test result for SARS-CoV-2.

### Sample collection and testing

Samples were collected by a clinical team at the study hospital, consisting of physicians, nurses, and biomedical laboratory scientists. Two nasopharyngeal samples were taken from the same nostril by trained staff from each participant. One sample was tested immediately at the hospital using the Standard Q SARS-CoV-2 Ag-RDT, with the result being interpreted according to the manufacture’s guidelines and recorded on a specific paper form. The second sample was transported in a viral transport medium to the zonal public health laboratory located within the premises of the Teaching Hospital for extraction and RT-PCR testing.

### Statistical methods

To determine sensitivity and specificity of the Standard Q SARS-CoV-2 Ag-RDT (with 95% CIs), results were compared to RT-PCR results from the same participant, as per Altman (17). A predefined subgroup analysis by symptoms presence was performed.

We used “R” version 4.0.3. (R Foundation for Statistical Computing, Vienna, Austria) to generate all analyses and plots.

### Ethical aspects

The evaluation protocol was approved by the Ethical Review Committee of the Tamale Teaching Hospital. The data used for the evaluation were routine data from the hospital services. Laboratory samples were anonymized and results could not be traced to individual participants. All participants provided verbal informed consent and there were no refusals.

## Results

A total of 198 participants were recruited for the study. For five participants, no RT-PCR results were available. Thus, 193 participants were included into the study for which both RT-PCR test and Ag-RDT results were available.

### Demographic characteristics

The median age was 35 years (range: 5 months to 93 years). 109 participants were male, 84 were female.

### Clinical characteristics

Table 1 shows the clinical characteristics of the study participants. 135 (70%) were symptomatic; 58 (30%) were asymptomatic. Cough (60%), fever (34%), general weakness (36%), rhinitis (36%) and headache (36%) were the main symptoms recorded. Ageusia and anosmia were recognized in only 16% and 19% of study participants respectively. Non-communicable diseases (NCDs) that are associated with severe COVID-19 were very rare (one case each of Diabetes mellitus, hypertension and cardiovascular disease), and none of the study participants was on medication for a chronic disease.

**Table 1:**
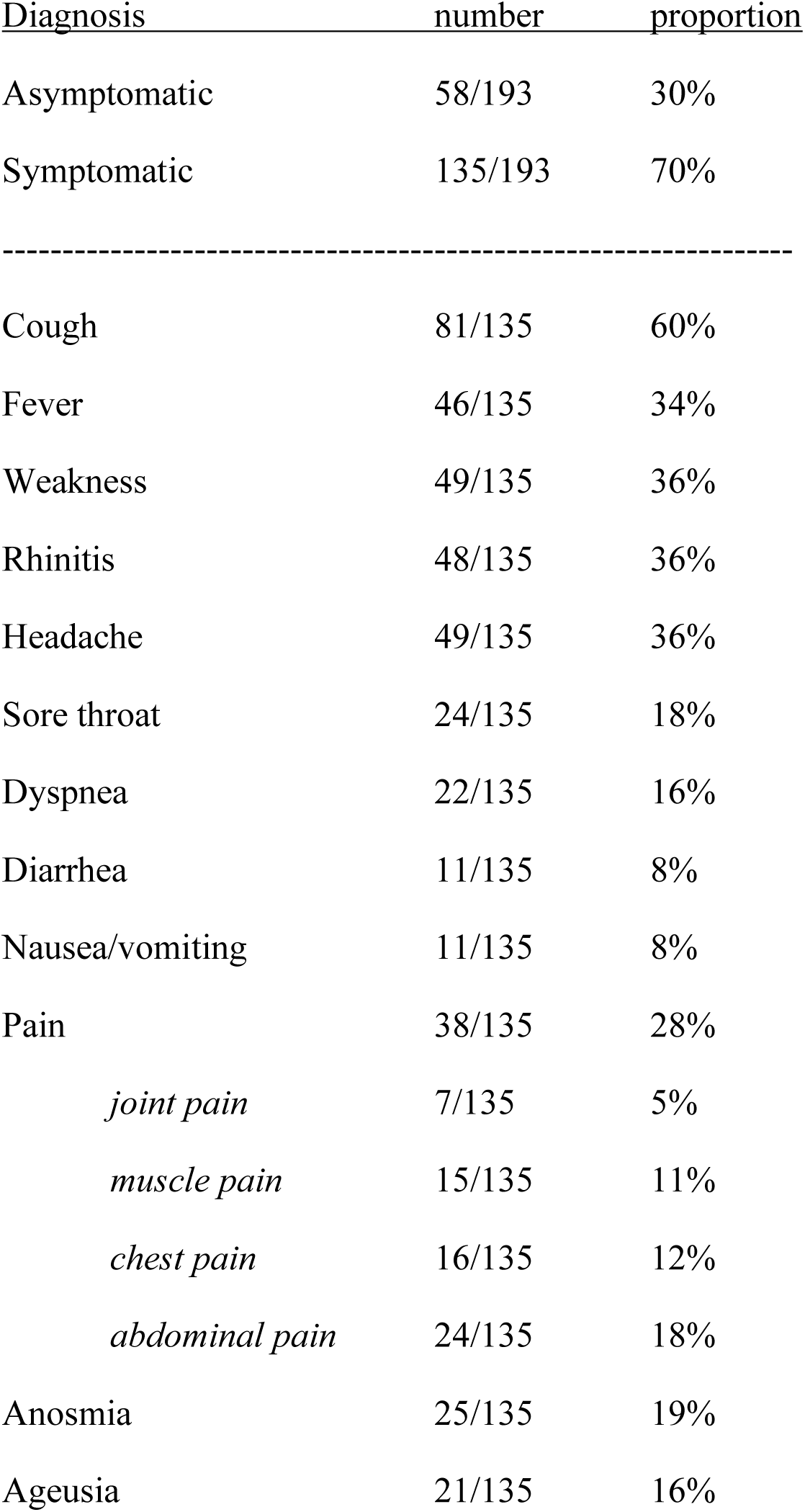
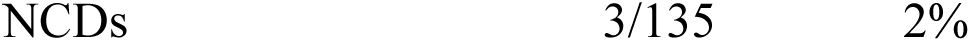
Clinical characteristics of study participants in Tamale Teaching Hospital, Ghana

| <u>Diagnosis</u> | <u>number</u> | <u>proportion</u> |
| --- | --- | --- |
| Asymptomatic | 58/193 | 30% |
| Symptomatic | 135/193 | 70% |
| ----- |  |  |
| Cough | 81/135 | 60% |
| Fever | 46/135 | 34% |
| Weakness | 49/135 | 36% |
| Rhinitis | 48/135 | 36% |
| Headache | 49/135 | 36% |
| Sore throat | 24/135 | 18% |
| Dyspnea | 22/135 | 16% |
| Diarrhea | 11/135 | 8% |
| Nausea/vomiting | 11/135 | 8% |
| Pain | 38/135 | 28% |
| *joint pain* | 7/135 | 5% |
| *muscle pain* | 15/135 | 11% |
| *chest pain* | 16/135 | 12% |
| *abdominal pain* | 24/135 | 18% |
| Anosmia | 25/135 | 19% |
| Ageusia | 21/135 | 16% |

### Test results

Table 2 and 3 show the results. A total of 42 (22%) study participants were RT-PCR positive, and 151 (78%) were RT-PCR negative. Of the 42 RT-PCR positives, 27 were Ag-RDT positive, resulting in a sensitivity of 64% (95% CI 49-79). 123 were identified Ag-RDT negative of the 151 RT-PCR negatives, resulting in a specificity of 81% (95% CI 75-87). The positive predictive value of the Ag-RDT in this population was 49% (95% CI 36-62) and the negative predictive value 89% (95% CI 84-94). The positive predictive accuracy (PPA) of the test was 28% and negative predictive accuracy (NPA) was 95%. Among the asymptomatic participants, there were 18 RT-PCR positives (31%) and 13 (72%) (95% CI 51-93) were detected by Ag-RDT. Only considering the symptomatic participants (24 RT-PCR positives, 18%), the sensitivity was 58% (95% CI 38-78). Regarding the cycle threshold (Ct) values of our participants who were PCR positive (40/42), none was <25; 8 (20%) had values between 25 and <30; 29 (72.5%) were between 30 and <35; and 3 (7.5%) were between 35 and <40.

**Table 2:**
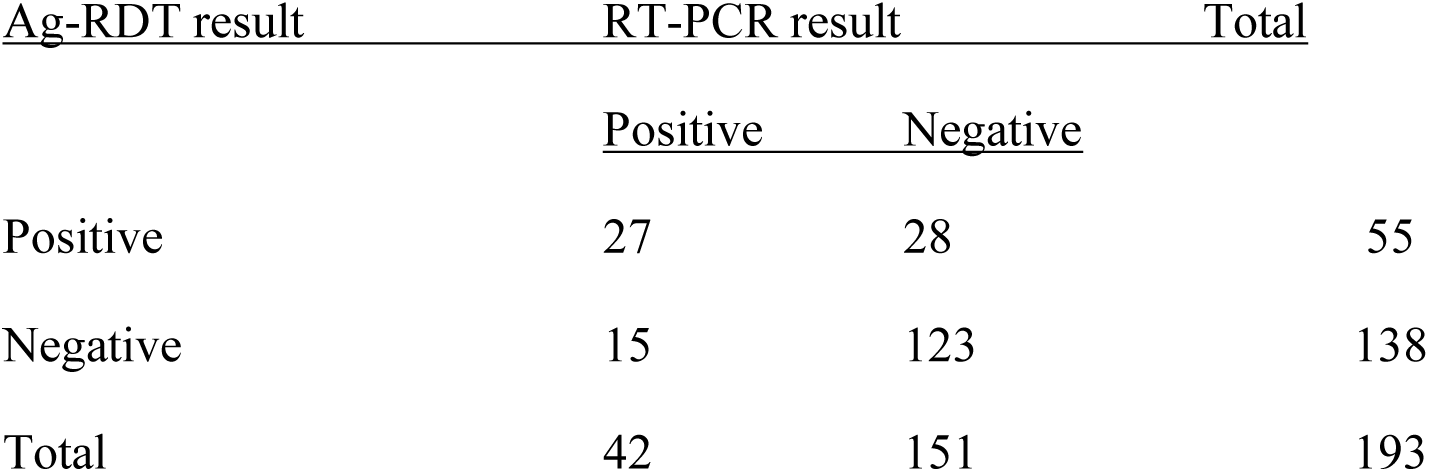
SARS-CoV-2 Ag-RDT test results in comparison to RT-PCR results in Tamale Teaching Hospital, Ghana

**Table 3:**
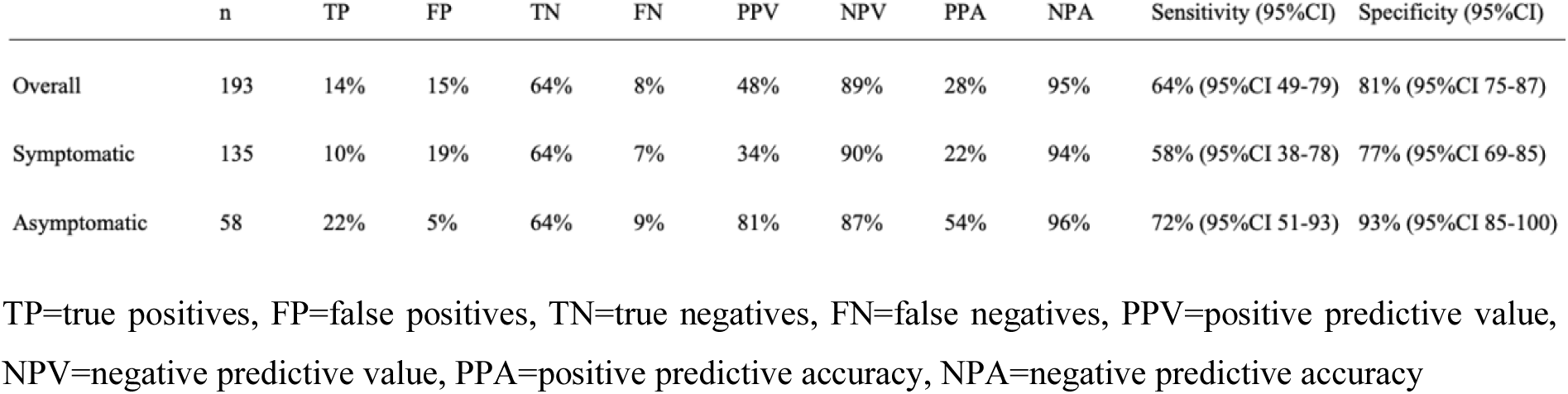
Subgroup analysis results of true/false positives and negatives in Tamale Teaching Hospital, Ghana

|  | n | TP | FP | TN | FN | PPV | NPV | PPA | NPA | Sensitivity (95%CI) | Specificity (95%CI) |
| --- | --- | --- | --- | --- | --- | --- | --- | --- | --- | --- | --- |
| Overall | 193 | 14% | 15% | 64% | 8% | 48% | 89% | 28% | 95% | 64% (95%CI 49-79) | 81% (95%CI 75-87) |
| Symptomatic | 135 | 10% | 19% | 64% | 7% | 34% | 90% | 22% | 94% | 58% (95%CI 38-78) | 77% (95%CI 69-85) |
| Asymptomatic | 58 | 22% | 5% | 64% | 9% | 81% | 87% | 54% | 96% | 72% (95%CI 51-93) | 93% (95%CI 85-100) |

TP=true positives, FP=false positives, TN=true negatives, FN=false negatives, PPV=positive predictive value, NPV=negative predictive value, PPA=positive predictive accuracy, NPA=negative predictive accuracy

## Discussion

Our study demonstrates a limited sensitivity (64%) and very low specificity (81%) for a WHO approved SARS-CoV-2 Ag-RDT, which is much below the performance demonstrated in systematic reviews (73,8%) and 99,7% respectively)(12) and even more so with the manufacturers data (sensitivity 76,6% and specificity 97,6%)(18).

Much of the data generated by the Standard Q Ag-RDT stem from high resource settings with temperate climates. There are limited data on evaluations of SARS-CoV-2 Ag-RDT available until very recently from lower-resourced settings with hotter climates (19). A study in Cameroon compared results from the SD Biosensor Standard Q SARS-CoV-2 Ag-RDT with RT-PCR results in asymptomatic and symptomatic adult participants from eight hospitals and demonstrated an overall sensitivity of 59% (95% CI 53-65), with the sensitivity increasing to 69% (95% CI 62-75) when only symptomatic participants were considered (20). Another study in Uganda compared results from another SARS-CoV-2 Ag-RDT with RT-PCR results in adult hospital patients and controls and demonstrated a sensitivity of 70% (95% CI 60-79) (21).

The sensitivity in our study (64% overall and 58% in symptomatic participants) is comparable to this data from Uganda and Cameroon. Interestingly, the sensitivity in asymptomatic participants in our study was higher than in symptomatic participants, but this did not reach statistical significance. A potential explanation could be that more of the asymptomatic patients were captured early in the disease when the viral load is high. The higher sensitivity in asymptomatic patients speaks more for participants related aspects affecting the sensitivity than operational or environmental factors, as those would be expected to affect performance in symptomatic and asymptomatic participants equally (22).

Specificity in our study (81%) is lower than that demonstrated in both Uganda and Cameroon (92%) (20, 21) and shows that under study conditions in SSA countries a much lower specificity than the WHO requested minimum specificity of 97% has to be expected. Possible explanations for the lower specificity in our study could be cross-reacting antibodies from previous infections or the environmental temperatures (24°-37°C) during the study period in the rooms (the general wards and the COVID-19 isolation center) where the tests were carried out. The Ct values for our patients, who were tested RT-PCR positive, were generally high. As reported in the results, none of the participants had Ct values < 25 and the majority of the values were between 30 to <35 (72.5%). This could have affected the performance of the RDTs as higher Ct values correspond with lower viral load and studies have shown that the performance of RDTs fall with increasing Ct values (12, 23). In addition, most testing occurred during the day when temperatures were high although direct sunlight was avoided. The manufacturer recommends a temperature of maximum 30°C and others have demonstrated that high temperatures above the manufacturer recommended targets negatively affect the test performance (22). While the study by Haage et al. demonstrated an effect of high temperatures primarily on sensitivity, others have shared the experience that it also substantially affects specificity (Denkinger personal communication). One way to minimize the effects of high temperatures on the test accuracy in these settings would be to conduct the tests in the mornings and evenings when the environmental temperature will be relatively lower. However, care must be taken not to negate the advantage of the RDT test, which is to provide rapid results to aid clinical decision making. Another important factor could be the high prevalence of other endemic infectious diseases, or others as yet unknown factors (4, 20).

Having Ag-RDTs with high specificity and sensitivity will be a key element for controlling the pandemic in settings like northern Ghana in particular as well as Ghana and SSA in general. This is due to the fact that the turnaround time for the gold standard test (PCR) was very long in both the first and second wave of the pandemic due to limited testing capacity. As a result, clinical decision making was delayed and increased the risk of further transmission. RDTs with their quick turnaround time could facilitate clinical decision making and contact screening of confirmed COVID-19 cases. The high negative predictive accuracy for the Ag-RDTs used in this study makes it a valuable tool for these purposes but all positive tests will require confirmation with RT-PCR test (24).

The limitations of this study are firstly that it was done in one center of one country only, thus limiting generalizability. Secondly, the overall sample size was small and further subgroup analyses to better understand the data were not possible. Thirdly, the temperature at time of testing was not recorded thus limiting a differentiated understanding of the impact of temperature. Another limitation is that the study did not include the duration between the onset of symptoms and the timing of the testing for the symptomatic participants. And while it is a limitation that the tests were not performed in manufacturer recommended temperature range, the strength of this study is, that it has been done under real life conditions in a high-temperature, limited resource country.

## Conclusion

This field evaluation of a standard SARS-CoV-2 Ag-RDT shows a rather low sensitivity and specificity. Given these findings, further studies are needed to assess the role of existing Ag-RDTs in high-temperature climates in Africa and around the world, where easy-to-use tests are urgently needed as RT-PCR testing is not widely available.

## Data Availability

The datasets generated during and/or analyzed during the current study are available from the corresponding author on reasonable request.

## Contributors

CMD, AJ, and AAM designed the study, OM wrote the first paper draft, AAM coordinated the field work, all authors contributed to the content of the paper.

## Acknowledgements

We thank the Covid-19 team of the *Tamale Teaching Hospital* for assisting with the sample collection, Dr. Kingsley Bimpong and Dr. Hadi Salawudini for assisting with data entry, and the staff of the *Zonal Public Health and Reference Laboratory*, Tamale, for assisting with the RT-PCR testing.

